# Estimation of the incubation period of SARS-CoV-2 in Vietnam

**DOI:** 10.1101/2020.05.09.20096800

**Authors:** Long V. Bui, Ha T. Nguyen, Hagai Levine, Ha N. Nguyen, Thu-Anh Nguyen, Thuy P. Nguyen, Truong T. Nguyen, Toan T. T. Do, Ngoc T. Pham, Hanh M. Bui

## Abstract

**Objective:** To estimate the incubation period of Vietnamese confirmed COVID-19 cases.

**Methods:** Only confirmed COVID-19 cases who are Vietnamese and locally infected with available data on date of symptom onset and clearly defined window of possible SARS-CoV-2 exposure were included. We used three parametric forms with Hamiltonian Monte Carlo method for Bayesian Inference to estimate incubation period for Vietnamese COVID-19 cases. Leave-one-out Information Criterion was used to assess the performance of three models.

**Results:** A total of 19 cases identified from 23 Jan 2020 to 13 April 2020 was included in our analysis. Average incubation periods estimated using different distribution model ranged from 6.0 days to 6.4 days with the Weibull distribution demonstrated the best fit to the data. The estimated mean of incubation period using Weibull distribution model was 6.4 days (95% credible interval (CrI): 4.89–8.5), standard deviation (SD) was 3.05 (95%CI 3.05–5.30), median was 5.6, ranges from 1.35 to 13.04 days (2.5th to 97.5th percentiles). Extreme estimation of incubation periods is within 14 days from possible infection.

**Conclusion:** This analysis provides evidence for an average incubation period for COVID-19 of approximately 6.4 days. Our findings support existing guidelines for 14 days of quarantine of persons potentially exposed to SARS-CoV-2. Although for extreme cases, the quarantine period should be extended up to three weeks.

## INTRODUCTION

COVID-19 caused by SAR-CoV-2 has been declared a global pandemic by WHO on March 11 2020 [1]. As a newly emerged disease, little have we known about the incubation period for COVID-19, for which patients have no clinical symptom but some infected persons can be contagious [2]. Understanding the distribution of incubation period is important to estimate the potential spreading of the SARS-CoV-2, and to determine an optimal duration of quarantine.

The incubation period is identified by the interval time between exposure to source of an infectious disease and the onset of the first clinical symptoms. As the incubation period is known, we would be able to model the current and future of the pandemic scale, and to evaluate the effectiveness of intervention strategies, therefore, would be able to act swiftly several intensive public health strategies for infectious diseases [3].

Since the first COVID-19 case found in Vietnam on 23 January 2020, as of 13 April 2020, there are 265 confirmed cases all over the country. Of those, two-thirds of the patients have recovered, and no death has been reported. The pandemic situation in Vietnam is significantly different from that in other countries. Vietnam has implemented strong measures against the SARS-CoV-2 transmission at an early stage. However, the country has limited resources, hence understanding the time of intensive monitoring will be helpful to plan for effective measures to minimize the risk for hidden SARS-CoV-2 infection. While there are several studies attempting to estimate the incubation period of SARS-CoV-2, to date, there is not yet a study estimating this parameter using data from Vietnam. This study aims to estimate the incubation period of Vietnamese confirmed COVID-19 cases.

## METHODS

### Data sources and collection

Data of all confirmed COVID-19 cases in Vietnam were made publicly available on the official website of Ministry of Health of Vietnam (ncov.moh.gov.vn). Case reports include patient number, travelling history, contact tracing and clusters, date of laboratory-confirmed, date of assertion, date and status of discharge. Additionally, the clusters of COVID-19 in a northern province of Vietnam have been described by Thanh et al [4]. Two researchers independently reviewed the full text of each case report of confirmed COVID-19 cases in Vietnam from 23 January 2020 to 13 April 2020, and entered data into a standardized case reporting form to establish a database for this study. Any discrepancies in data extraction were resolved by discussion between two researchers and facilitated by a third researcher to reach consensus.

To assure the reliability of analysis, we selected only confirmed COVID-19 cases who are Vietnamese and locally infected with available data on date of symptom onset (including fever, cough, and shortness of breath) and clearly defined window of possible SARS-CoV-2 exposure. This window period is defined as the date range between the earliest possible exposure, which is the first contact with confirmed cases and the latest exposure, which is the most recent contact with confirmed cases in the clusters.

### Statistical analysis

We assumed that the moment of infection occurred between the interval of possible SARS-CoV-2 exposure. The distribution of incubation period was estimated using maximum likelihood where the likelihood function of each case in the dataset was a single interval-censored by three parametric forms with Hamiltonian Monte Carlo method for Bayesian Inference: Weibull distribution, the Gamma distribution and the Lognormal distribution. Non-informative positive prior for the parameters of the three distributions were specified. Leave-one-out Information Criterion (LooIC) was used to assess the performance of three models. The differences of LooIC larger than two were considered as statistically significant [5]. Mean, median and posterior Credible Interval (CI) for each distribution were also estimated. Statistical analysis was conducted by rstan [6] in R 3.6.4 [7]. Data for analysis and R codes are available on public repository https://github.com/longbui/Covid19IncubVN

## RESULTS

From 23 Jan 2020 to 13 Apr 2020, a total of 265 positive-confirmed SARS-CoV-2 was reported in Vietnam. Of those, 163 cases were identified as imported, 102 were locally infected. There were 19 confirmed COVID-19 cases found between 23 January to 13 April 2020, who met the inclusion criteria. These include being Vietnamese nationality, locally infected with SARS-CoV-2, and had completed epidemiological data on exposure interval and date of symptom onset. Of these cases, 7 were male and 12 were female. The median age was 37 (interquartile range: 25 – 50).

Average incubation periods estimated using different distribution model ranged from 6.0 days to 6.4 days (Table 1). According to the LooIc of proposed models, the Weibull distribution demonstrated the best fit to the data. The estimated mean of the incubation period using Weibull distribution model was 6.4 days (95% CI 4.89 –8.5), the standard deviation (SD) was 3.05 (95%CI 3.05 – 5.30). The Gamma distribution model fitted significantly poorer than the Weibull model. The estimated mean of the incubation period of this model was 6.07 days (95% CI: 4.64 – 8.00), with the SD of 2.90 days (95% CI 1.91 – 4.80), The lognormal distribution showed the poorest fit to the data, the incubation period was estimated to be 6.4 (95% CI 4.6 – 10.2).

**Table 1.** Mean and SD of the estimated incubation period for confirmed Vietnamese COVID-19 cases between 23 Jan 2020 to 13 Apr 2020

| Distribution | Mean (days) |  | SD (days) |  | LooIC |
| --- | --- | --- | --- | --- | --- |
|  | Estimate | 95% CI | Estimate | 95% CI |  |
| Weibull | 6.4 | 4.9–8.5 | 3.0 | 2.0–5.4 | 103.9 |
| Gamma | 6.1 | 4.7–8.0 | 2.9 | 1.9–4.8 | 106.2 |
| Lognormal | 6.4 | 4.6–10.2 | 5.8 | 3.5–11.2 | 106.7 |

Estimation of median of incubation period using different models are presented in Table 2. Figure 1 shows the estimation distribution of incubation periods using Weibull model, with estimated median of 6.1 days, ranging from 1.4 to 13.0 days (2.5^th^ to 97.5^th^ percentiles). Detail estimated possible moment of infection with Weibull distribution is illustrated in Figure 2. The results suggest a large variation in incubation periods among patients but it was most likely to fall in the period within 14 days of infection.

**Figure 1.**
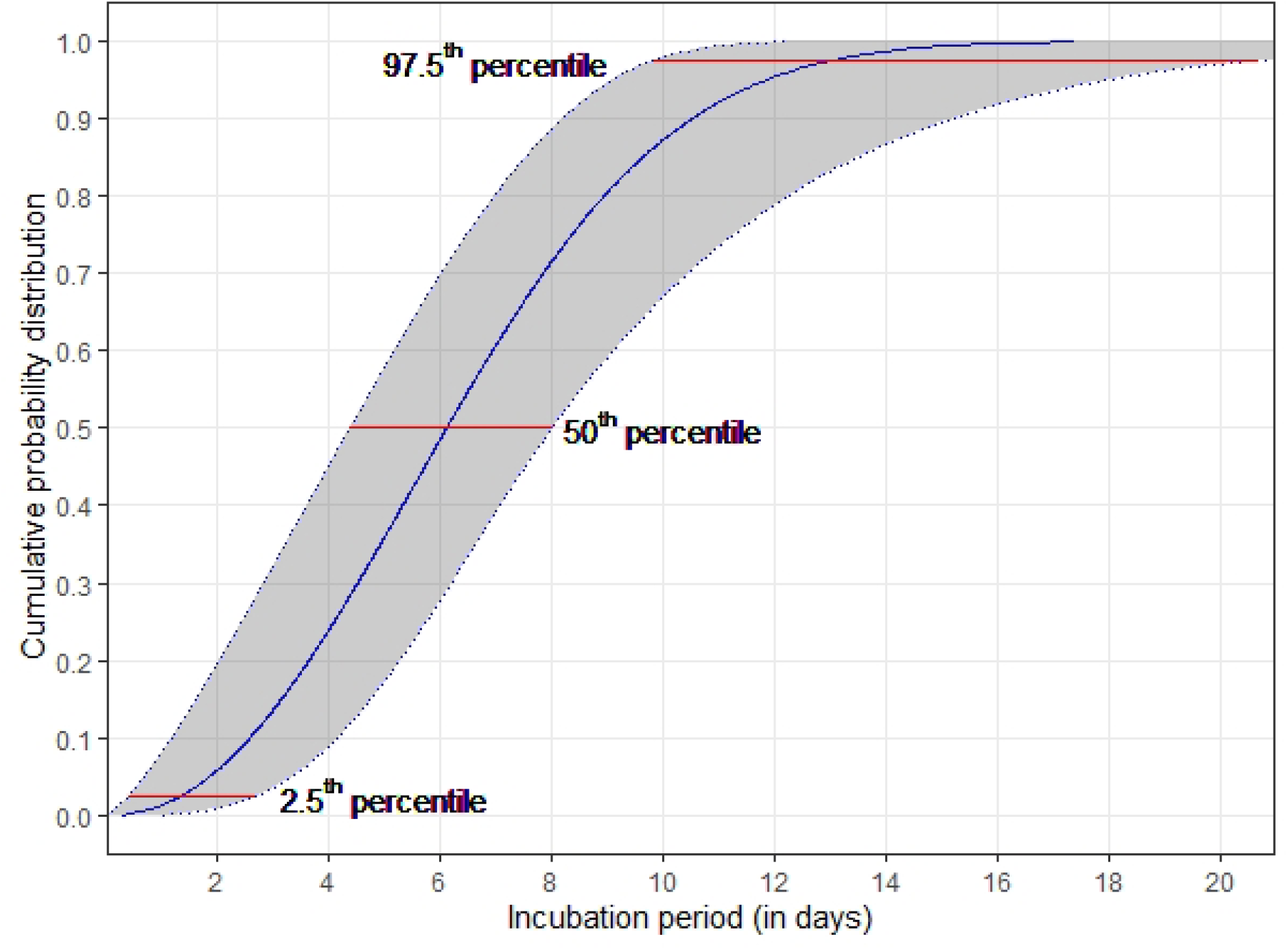
The cumulative density function of the estimated Weibull distribution for incubation period of Vietnamese confirmed COVID-19 cases from 23 Jan 2020 to 13 Apr 2020

**Figure 2.**
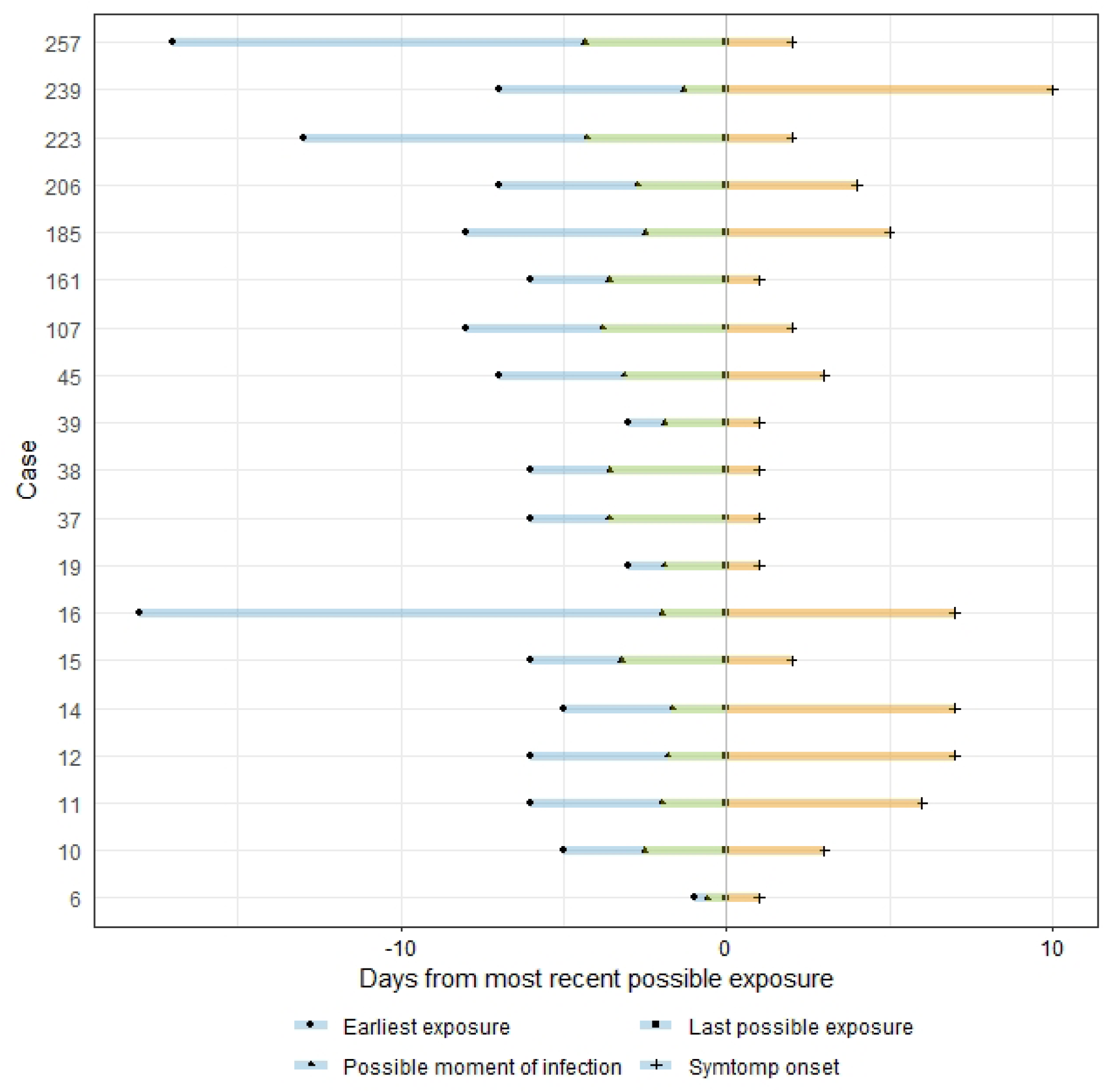
Estimated possible moment of infection of Vietnamese confirmed COVID-19 cases from 23 Jan 2020 to 13 Apr 2020 with Weibull distribution *(Cases were numbered by the order of official assertion of confirmed cases on the website of the Ministry of Health of Vietnam)*

**Table 2.** Percentiles of the estimated incubation period for confirmed Vietnamese COVID-19 cases between 23 Jan 2020 to 13 Apr 2020 with Weibull, Gamma and Lognormal distribution

| Percentiles | Incubation period distribution (days) |  |  |  |  |  |
| --- | --- | --- | --- | --- | --- | --- |
|  | Weibull |  | Gamma |  | Lognormal |  |
|  | Estimate | 95% CI | Estimate | 95% CI | Estimate | 95% CI |
| 2.5<sup>th</sup> | 1.4 | 0.4–2.7 | 1.8 | 0.7–33.0 | 1.5 | 0.6–2.5 |
| 5<sup>th</sup> | 1.9 | 0.7–3.4 | 1.0 | 2.2–3.4 | 1.8 | 2.0–3.5 |
| 50<sup>th</sup> | 6.1 | 4.4–8.0 | 5.6 | 4.2–7.3 | 5.2 | 3.6–7.3 |
| 95<sup>th</sup> | 11.9 | 9.10–12.0 | 11.5 | 8.6–16.7 | 14.9 | 9.7–30.2 |
| 97.5<sup>th</sup> | 13.0 | 9.1–20.7 | 13.0 | 9.6–19.4 | 18.2 | 11.3–40.8 |
| 99<sup>th</sup> | 14.4 | 10.6–24.0 | 14.7 | 10.7–22.8 | 23.0 | 14.4–58.0 |

## DISCUSSION

We estimated the incubation period of Vietnamese confirmed COVID-19 cases from public reported data with three parametric models, including Weibull, Gamma and Lognornal distribution. The Weibull distribution proved to be best fit to the data. Our estimation results are similar or higher than most results from published literature (Table 3). Our estimation of 6.4 days for mean incubation period is similar to the estimation of Backer et al [8]. The estimated mean in our study is higher in comparison with that of other studies [9-12], which ranged from 4.9 to 5.6 days. Only one study by Leung [13] estimated longer incubation periods than our estimation among local residents of Wuhan. When comparing the estimated median and its range with other study, we saw a similar trend where our estimations are similar or higher than most studies where median estimation was available [8, 10-12]. Nonetheless, extreme estimation of incubation periods (95^th^ or 97.5^th^ percentiles) in studies are well within 14 days from possible infection [8, 10-13].

**Table 3.** Comparison of estimated incubation periods for SARS-COV-2

| Author | Distribution | N | Mean and/or median (days) with 95%CI | Plausible range (days) with 95%CI |
| --- | --- | --- | --- | --- |
| Our study | Weibull | 19 | Mean 6.4 (4.9 - 8.2)<br>Median 6.1 (4.4 - 8.0) | 1.4 - 13.0 (2.5<sup>th</sup> to 97.5<sup>th</sup> percentile) |
| Backer et al.[8] | Weibull | 88 | Mean 6.4 (5.6 - 7.7)<br>Median 6.4 (5.5 - 7.5) | 2.1-11.1 days (2.5 to 97.5 percentile) |
| Lauer et al.[10] | Lognormal | 181 | Median 5.2 (4.4 - 6.0) | 2.2-11.5 (2.5<sup>th</sup> to 97.5<sup>th</sup> percentile) |
| Linton et al.[12] | Lognormal | 58<br>(excluding Wuhan residents) | *Non-truncated*<br>Mean 5.0 (4.2 - 6.0),<br>Median 4.3 (3.5 - 5.1)<br>*Right-truncated*<br>Mean 5.6 (4.4 - 7.4)<br>Median 4.6 (3.7 - 5.7) | 95<sup>th</sup> percentile for non- truncated data: 10.6 (8.5 - 14.1)<br>95<sup>th</sup> percentile for right-truncated: 12.3 (9.1-19.8) |
|  | Lognormal | 152<br>(including Wuhan residents) | Nontruncated:<br>Mean 5.6 (5.0 - 6.3),<br>Median 5.0 (95%: 5.0 - 6.3) | 95<sup>th</sup> percentile for non-truncated data 10.8 (95% CI: 9.3 - 12.9) |
| Li et al.[11] | Lognormal | 10 | Mean 5.2 (4.1 - 7.0), | 95<sup>th</sup> percentile 12.5 (9.2 - 18) |
| Leung [13] | Weibull | 175<br>(Travelers to Hubei) | Mean 1.8 (1.0 - 2.7) | 95<sup>th</sup> percentile 3.2 (1.0 - 3.8) |
|  | Weibull | 175 (Non-travelers) | Mean 7.2 (7.1 - 8.4) | 95<sup>th</sup> percentile 14.6 (12.1 - 17.1) |
| Jiang et al.[9] |  | 50 | Mean 4.9 (4.4 - 5.5) |  |

The estimated mean of incubation period for 19 Vietnamese confirmed COVID-19 cases using Weibull distribution model is higher than that of SARS in Hong Kong and Beijing [14] and in MERS [15, 16]. Hence, mathematical models of COVID-19 should not be fitted with incubation period of MERS or SARS.

The variation of incubation period of SARS-CoV-2 could be explained by the difference on the strain of the virus, the biological variations of population and the control measures of certain country. It is suggested that any interpretation of results on incubation time of SARS-CoV-2 is strongly dependent on the selected data sources [9]. In Vietnam, the key COVID-19 control strategy has focused on active case finding, early testing, treating and strictly isolating cases, which has been considered as effective in finding people with SARS-CoV-2 positive and providing treatment prior to clinical symptoms presentation. It is also a possible circumstance that people with suggestive symptoms of respiratory diseases may take drugs before seeking diagnosis and treatment.

In this study, the plausible range of incubation period estimated using the Weibull distribution supports the importance to isolate confirmed and suspected cases, and close contacts for 14 days after exposure. However, we recommend the policy makers to consider the upper bound of this range (97.5th percentile 13.0, CI 10.6–20.7). As such, the quarantine period should be extended up to three weeks.

Our study has several strengths, including being the first effort to estimate SAR-CoV-2 incubation period using data from Vietnamese locally transmitted cases. We also used 3 different models for estimation and identified the model that best fitted with our data. However, there are potential limitations in our study. First, not all confirmed COVID-19 cases have completed epidemiological data on exposure interval and date of symptom onset available on public government reports. Second, the number of patients included in the analysis is small, possibly explaining the wider range of estimated incubation period. This highlights the need for data standardization for further studies on COVID-19 and the importance of publishing data for the scientific and public health community [9].

This study may contribute to the effort of COVID-19 control effort in Vietnam and elsewhere by providing an informed estimate of the incubation period. This is a key variable needed for modelling the spread of SARS-CoV-2, for informed decision-making throughout the pandemic. Similar methodology could be used to estimate the incubation period for other countries and diseases.

## Data Availability

Data for analysis and R codes are available on public repository https://github.com/longbui/Covid19IncubVN

## Author Contributions

HMB, TAN, HTN, and LVB had full access to all of the data in the study and takes responsibility for the integrity of the data and the accuracy of the data analysis. ATTN, HL, HNN, TTTD, TTN, TPN, TNP and HMB contributed substantially to the study design, data analysis and interpretation, and the writing of the manuscript.

### Funding

The author(s) received no specific funding for this work.

### Disclaimer

The views and opinions expressed in this article are those of the authors and do not necessarily reflect the official policy or position of any agency of the government.

## References

1. WHO. WHO characterizes COVID-19 as a pandemic. Rolling updates on coronavirus disease (COVID-19) 2020 [cited 2020 Apr 20]; Available from: https://www.who.int/emergencies/diseases/novel-coronavirus-2019/events-as-they-happen.

2. WHO, Coronavirus disease 2019 (COVID-19) Situation Report - 73, in Coronavirus disease 2019 (COVID-19) Situation Report, WHO, Editor. 2020, WHO: Geneva.

3. Nishiura, H., Early efforts in modeling the incubation period of infectious diseases with an acute course of illness. Emerging Themes in Epidemiology, 2007. 4(1): p. 2.

4. Thanh, H.N., et al., Outbreak investigation for COVID-19 in northern Vietnam. The Lancet Infectious Diseases, 2020. 0(0).

5. Vehtari, A., A. Gelman, and J. Gabry, Practical Bayesian model evaluation using leave-one-out cross-validation and WAIC. Statistics and Computing, 2017. 27(5): p. 1413-1432.

6. Stan Development Team, RStan: the R interface to Stan. 2020.

7. Team, R.C., R: A Language and Environment for Statistical Computing. 2017, Vienna, Austria.

8. Backer, J.A., D. Klinkenberg, and J. Wallinga, Incubation period of 2019 novel coronavirus (2019-nCoV) infections among travellers from Wuhan, China, 20–28 January 2020. Eurosurveillance, 2020. 25(5): p. 2000062.

9. Jiang, X., S. Rayner, and M.-H. Luo, Does SARS-CoV-2 has a longer incubation period than SaRS and MERS? Journal of Medical Virology, 2020. 92(5): p. 476–478.

10. Lauer, S.A., et al., The Incubation Period of Coronavirus Disease 2019 (COVID-19) From Publicly Reported Confirmed Cases: Estimation and Application. Annals of Internal Medicine, 2020.

11. Li, Q., et al., Early Transmission Dynamics in Wuhan, China, of Novel Coronavirus–Infected Pneumonia. New England Journal of Medicine, 2020. 382(13): p. 1199-1207.

12. Linton, N.M., et al., Incubation Period and Other Epidemiological Characteristics of 2019 Novel Coronavirus Infections with Right Truncation: A Statistical Analysis of Publicly Available Case Data. Journal of Clinical Medicine, 2020. 9(2): p. 538.

13. Leung, C., The difference in the incubation period of 2019 novel coronavirus (SARS-CoV-2) infection between travelers to Hubei and non-travelers: The need of a longer quarantine period. Infection Control and Hospital Epidemiology, 2020: p. 1–3.

14. Lau, E.H.Y., et al., A comparative epidemiologic analysis of SARS in Hong Kong, Beijing and Taiwan. BMC Infectious Diseases, 2010. 10(1): p. 50.

15. Assiri, A., et al., Hospital Outbreak of Middle East Respiratory Syndrome Coronavirus, in http://dx.doi.ora/10.1056/NEJMoa1306742. 2013.

16. Cauchemez, S., et al., Middle East respiratory syndrome coronavirus: quantification of the extent of the epidemic, surveillance biases, and transmissibility. The Lancet Infectious Diseases, 2014. 14(1): p. 50–56.

